# Racial Disparities in Hesitancy and Utilization of Monoclonal Antibody Infusion Treatment of COVID-19

**DOI:** 10.1101/2022.07.07.22277360

**Authors:** Yahya Shaikh, Ishaan Gupta, Sophia Purekal, Mary Jane E. Vaeth, Maisha Foyez, Charles D. Callahan, Maryam Elhabashy, James R. Ficke, Albert W. Wu, Paul G. Auwaerter, Melinda E. Kantsiper, Zishan K. Siddiqui

## Abstract

**Background and Methods:** We conducted a single center cross-sectional study to investigate racial disparities in the hesitancy and utilization of monoclonal antibody (mAb) treatment of COVID-19 among treatment eligible patients who were referred to the infusion center between January 4, 2021 and May 14, 2021.

**Results:** Among the 2,406 eligible participants, African Americans were significantly more likely to underutilize mAb treatment (OR 1.8; 95% CI 1.5-2.1) and miss treatment opportunities due to monoclonal hesitancy (OR 1.7, 95% CI 1.3-2.1).

**Conclusion:** Addressing racial disparities in mAb delivery is an opportunity to bridge the racial inequities in COVID-19 care.

## Background

Since the start of the COVID-19 pandemic, members of racial and ethnic minorities have borne a disproportionate burden of disease with disparities in infection rates, hospitalization, and death.^1,2^ Further, differences have also been seen in preventive measures, with African-Americans expressing a rate of vaccine hesitancy three times greater than whites.^3^ Targeted interventions to address barriers and improve equity in COVID-19 outcomes have included uptake of vaccines.^4,5^ However, whether access to highly effective monoclonal antibody (mAb) infusions for the treatment of COVID-19 face similar issues is unknown.

Three anti-SARS-CoV-2 monoclonal antibody products have received Emergency Use Authorizations (EUAs) from the Food and Drug Administration (FDA) to treat mild to moderate COVID-19 in non-hospitalized patients at high risk for progressing to severe disease and/or hospitalization. These have been shown to improve outcomes.^6^ We investigated if there is evidence for racial disparities in utilization of mAb and treatment hesitancy after referral to a high-volume infusion site.

## Methods

This was a cross-sectional analysis of patients seen at the mAb infusion site linked to the Baltimore Convention Center Field Hospital (BCCFH), Maryland’s largest infusion site. The operational structure for BCCFH has been described.^7^ All patients referred to the center from January 4^th^, 2021 to May 14^th^, 2021, who were eligible to receive mAb treatment based on their disease severity and risk factors, were included in the study. Patients were referred by community providers using a web-based form that automatically populated the scheduling spreadsheet. Treatment was provided with no out-of-pocket payment irrespective of insurance status. Free transportation was offered to and from the infusion center. Patients outside of the 10-day eligibility period were excluded and marked as “Timed Out.” Attempts were made to contact eligible patients within 24 hours to schedule a treatment. Additional attempts were made, and the referring provider was contacted via automated email after two failed attempts. The disposition status of the patient was documented after each contact attempt (e.g., “Unable to be Reached,” “Declined,” “Cancelled,” “Hospitalized,” “Treated Elsewhere,” “Scheduled”). The scheduled patient’s status was updated on the planned infusion date (“No-Show,” “ED-transfer after Arrival,” “Treated”). Final disposition status of “Refused Treatment” or “Cancelled” was categorized as mAb hesitancy partly because further inquiry often revealed doubts about the treatment. Treatment at BCCFH or elsewhere was categorized “Appropriate Utilization.” All other disposition statuses of referred patients and eligible patients that did not lead to treatment were considered “Underutilization.”

Racial disparities were assessed using logistic regression models to compare odds by race for treatment utilization, final disposition status, overall underutilization, and disposition consistent with mAb hesitancy. All analyses were conducted as both unadjusted models and models adjusted for gender, age, and poverty rate using the patient’s zip code of residence as a surrogate for socioeconomic status derived from the American Community Survey.^8^ All statistical testing was 2-tailed with an α□=□.05 using SAS Studio Software, Version 3.8 of SAS (SAS Institute Inc., Cary, NC, USA). The study was approved by local institutional review board. The dataset is not publicly available.

## Results

Between 1/04/2021 and 05/14/2021, 2406 eligible patients were referred, of which 57.4% (1382) were white, 35% (841) were African American, 3.4% were Latinx, 3.1% Asian, with Native Americans, Pacific Islanders, and self-identified categories making up less than 1%. African American patients were younger, with a mean age of 54.6 (95% CI, 53.6-55.6) vs. 61.3 years (95% CI, 60.5-62.1) for whites, and were more often from neighborhoods with higher levels of poverty (Table 1). Overall, 69.5% (95% CI, 67.6-71.3) of referred patients were treated at BCCFH, with a greater proportion of whites receiving mAb (74% [95% CI, 71.7-76.3]) vs African American (63.8% [95%CI, 60.5-67.0]).

**Table 1.** Patients Demographics.

|  | **Total (%)**<br>**N=2406** | **African-American**<br>**(%)**<br>**N=841** | **White (%)**<br>**N=1382** | **p-value** |
| --- | --- | --- | --- | --- |
| **Age (years old)** | 58.6 (58.0-59.2) | 54.6 (53.6-55.6) | 61.3 (60.5-62.1) | <0.0001 |
| **Age Category (years old)** |  |  |  |  |
| <55 | 33.1 (31.2-35.0) | 44.2 (40.8,47.5) | 25.3 (23.0,27.6) | <0.0001 |
| >= 55 | 66.9 (65.0-68.8) | 55.8 (52.5,59.2) | 74.7 (72.4,77.0) |  |
| **Gender** |  |  |  |  |
| Female | 56.9 (54.9-58.9) | 66.6 (63.4-69.8) | 51.7 (49.0-54.3) | <0.0001 |
| Male | 43.1 (41.1-45.1) | 33.4 (30.2-36.6) | 48.3 (45.7-51.0) |  |
| **Rental Housing in**<br>**Patient's Zipcode** |  |  |  |  |
| 0 – 20% | 27.9 (26.1-29.7) | 7.8 (6.0-9.7) | 40.5 (37.9-43.1) | <0.0001 |
| 20 – 40% | 42.5 (40.5-44.5) | 40.4 (37.1-43.7) | 42.8 (40.2-45.4) |  |
| 40%+ | 29.6 (27.8-31.4) | 51.8 (48.4-55.2) | 16.6 (14.7-18.6) |  |
| **Poverty Rate in Patient's**<br>**Zipcode** |  |  |  |  |
| 0 - 10% | 64.5 (62.6-66.4) | 39.2 (35.9-42.5) | 78.8 (76.6-81.0) | <0.0001 |
| 10 - 20% | 23.0 (21.3-24.7) | 31.4 (28.2-34.5) | 18.0 (16.0-20.0) |  |
| 20+ % | 12.5 (11.2-13.8) | 29.5 (26.4-32.5) | 3.2 (2.3-4.1) |  |
| **Baltimore City Residence** | 39.8 (37.8-41.7) | 62.6 (59.3-65.9) | 27.3 (24.9-29.6) | <0.0001 |

Among referred patients, 12.6% (95% CI, 10.3-14.8) of African Americans and 8.0% (95% CI, 6.5-9.4) of whites refused treatment; and 8.7% (95% CI, 6.8-10.6) of African Americans and 4.5% of whites (95% CI, 3.4-5.6) were unreachable. (Figure 1) Overall, mAb hesitancy accounted for 48.0% (95% CI, 44.3-51.7) of underutilization.

**Figure 1.**
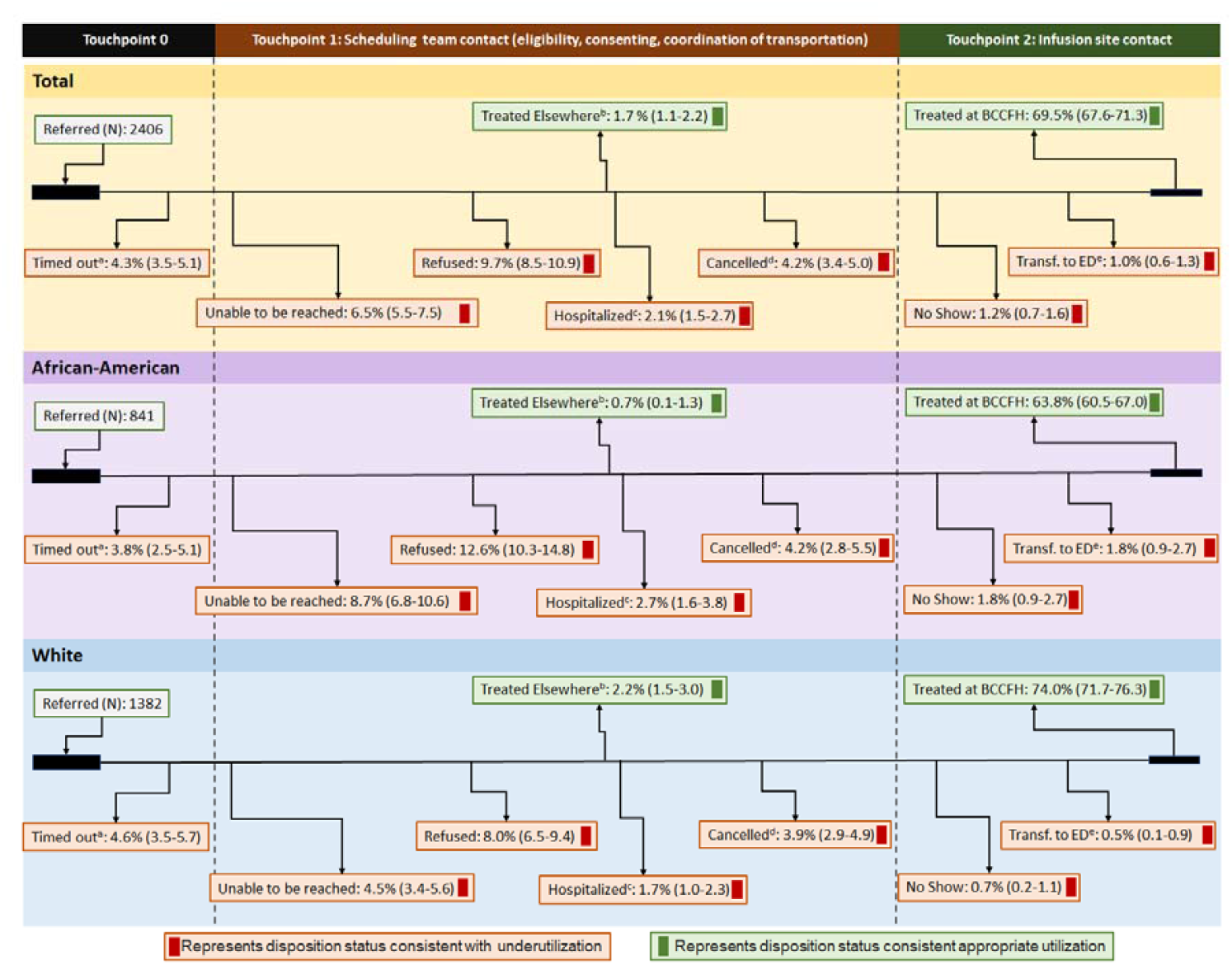
Distribution of Patients Final Disposition Status Between Referral and Treatment by Race. a. Timed out: patients past 10-day window of eligibility after symptom onset at the time of referral b. Hospitalized: patients hospitalized when scheduling call was made c. Treated elsewhere: on initial call declined appointment because treated elsewhere OR called to cancel an appointment at BCCFH because treated elsewhere d. Cancelled: patients called to cancel the appointment without being scheduled elsewhere e. Transferred to ED: Patient unstable on arrival to infusion site and transferred to ED prior to any treatment

Compared to whites, African-Americans had 3.5 (95% CI, 1.4-8.8) times greater odds of being transferred to the Emergency Department on arrival to infusion site; 2.0 times greater odds (95% CI, 1.4-2.9) of being unreachable during the 10-day eligibility window for mAb treatment; and 1.7 times greater odds (95% CI, 1.3-2.2) of refusing treatment.

African Americans had greater odds of under-utilizing mAb infusion treatment (1.8; 95% CI: 1.5, 2.1) and missing treatment opportunities because of monoclonal hesitancy (1.7; 95% CI: 1.3-2.1). Whites had greater odds of being treated at BCCFH (OR 1.6; 95% CI, 1.3-1.9) or another infusion site (3.2; 95% CI, 1.3-7.7). The results were similar after adjusting for age, gender and socioeconomic status (Table 2).

**Table 2.**
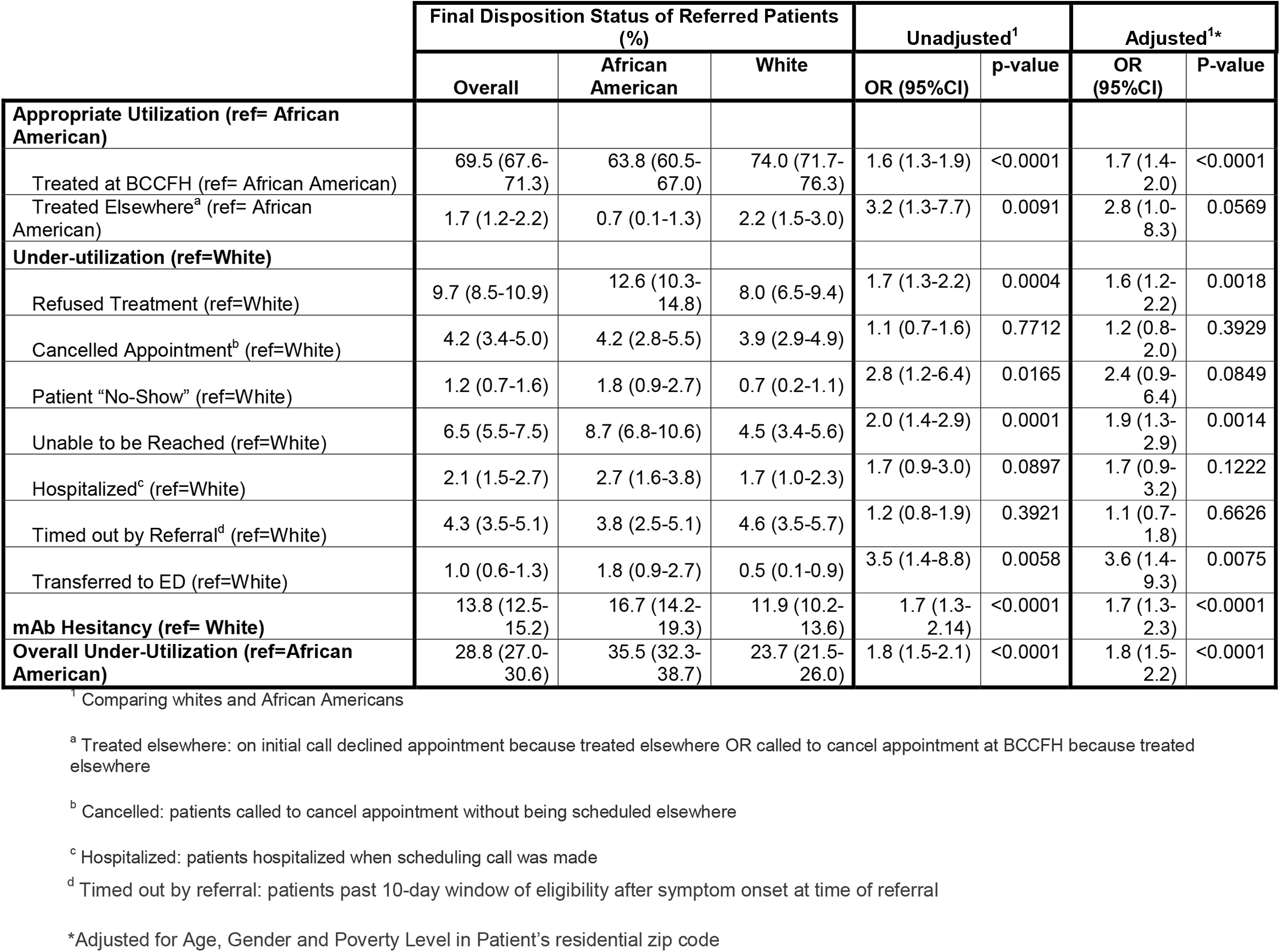
Comparison of Disposition Status by Race.

## Discussion

This study of patient utilization patterns after referral for mAb reveals disparities between whites and African-Americans. This occurred despite efforts to mitigate barriers to utilization, including free transportation and zero out-of-pocket expenses. African Americans were more likely to underutilize mAb after referral. Higher rates of mAb hesitancy appear to be the main reason for underutilization. Additional factors included delays due to disease severity, requiring emergency department transfer on infusion date, inability to follow-up reliably as demonstrated by no-show rates, and failure to be contacted.

Compared to whites, African Americans had higher odds of underutilization and mAb hesitancy as a final disposition status. This suggests that race is a strong driver of underutilization and hesitancy in the overall referred population. Hesitancy has been cited as a reason for the low uptake of mAb treatment.^9^ The higher prevalence of mAb hesitancy among African Americans in our population parallels patterns in COVID-19 vaccine hesitancy.^10^ A significant cause of vaccine hesitancy for African-Americans relates to concerns about safety and efficacy,^11^ worry that treatment may be harmful with a lack of trust in COVID treatment, and distrust related to unethical historical research practices among the Blacks.^12^ Evidence-based approaches have been used to address social, individual, and structural issues related to vaccine hesitancy.^4,5^ A similar approach will be needed to uncover reasons for mAb hesitancy and overcome it. Hesitancy to mAb may not be as entrenched as vaccines, but this treatment may also have a different risk-benefit profile. Delays in care and loss to follow-up may be related to economic barriers, patient perceptions of their needs, and fragmentation of care. These patients may benefit if efforts to achieve equity in Covid care were expanded include to mAb treatment.

As a single-site study, the generalization of these results is limited. In addition, we may have missed patients who utilized services elsewhere. However, our site provided over a third of infusions in the state, and our study suggests that African Americans were less like to use services elsewhere. In conclusion, African Americans disproportionately underutilize anti-SARS-CoV-2 monoclonal antibody treatment despite referral. Hesitancy about mAb treatment may be the primary reason. There may be a similar problem in the community leading to non-referral. Efforts to better understand mAb hesitancy and other barriers, and proactive outreach similar to those employed for Covid-19 vaccination, may be needed to mitigate barriers for underserved communities and high-risk patients.

## Data Availability

Data is not available.

## Authorship confirmation statement

All persons listed as authors meet the authorship criteria and all authors have made significant contribution in conception, design, analysis, writing and revision of the manuscript. All authors have read and approved the manuscript.

## Acknowledgements

None

## Meeting Presentations

This work has not been presented at any meeting.

## References

1. Romano SD, Blackstock AJ, Taylor EV, et al. Trends in Racial and Ethnic Disparities in COVID-19 Hospitalizations, by Region - United States, March-December 2020. MMWR. Morbidity and mortality weekly report 2021;70:560–5.

2. Yaya S, Yeboah H, Charles CH, Otu A, Labonte R. Ethnic and racial disparities in COVID-19-related deaths: counting the trees, hiding the forest. BMJ global health 2020;5:e002913.

3. Nguyen LH, Joshi AD, Drew DA, et al. Racial and ethnic differences in COVID-19 vaccine hesitancy and uptake. medRxiv : the preprint server for health sciences 2021.

4. Grumbach K, Carson M, Harris OO. Achieving Racial and Ethnic Equity in COVID-19 Vaccination. JAMA health forum 2021;2:e211724.

5. Evans A, Webster J, Flores G. Partnering With the Faith-Based Community to Address Disparities in COVID-19 Vaccination Rates and Outcomes Among US Black and Latino Populations. JAMA : the journal of the American Medical Association 2021;326:609–10.

6. National Institute of Health. Anti-SARS-CoV-2 Monoclonal Antibodies. Available at: https://www.covid19treatmentguidelines.nih.gov/therapies/anti-sars-cov-2-antibody-products/anti-sars-cov-2-monoclonal-antibodies. Accessed 10/, 2021.

7. Jones JA, Siddiqui ZK, Callahan C, et al. Infection Prevention Considerations for a Multi-Mission Convention Center Field Hospital in Baltimore, Maryland, During the COVID-19 Pandemic. Disaster medicine and public health preparedness 2021:1–8.

8. U.S. Census Bureau; American Community Survey, 2015-2019 American Community Survey 5-Year Estimates, Table S1701. Retrieved from https://data.census.gov/cedsci/table?q=poverty&g=0400000US24,24%248600000&tid=ACSST5Y2019.S1701; (11 October 2021).

9. Anderson TS, O’Donoghue AL, Dechen T, Mechanic O, Stevens JP. Uptake of Outpatient Monoclonal Antibody Treatments for COVID-19 in the United States: a Cross-Sectional Analysis. Journal of general internal medicine : JGIM 2021.

10. Momplaisir FM, Kuter BJ, Ghadimi F, et al. Racial/Ethnic Differences in COVID-19 Vaccine Hesitancy Among Health Care Workers in 2 Large Academic Hospitals. JAMA Network Open 2021;4:e2121931.

11. Callaghan T, Moghtaderi A, Lueck JA, et al. Correlates and Disparities of COVID-19 Vaccine Hesitancy. SSRN Electronic Journal.

12. Momplaisir F, Haynes N, Nkwihoreze H, Nelson M, Werner RM, Jemmott J. Understanding Drivers of COVID-19 Vaccine Hesitancy Among Blacks. Clinical infectious diseases 2021

